# Development and validation of a multimodal neuroimaging biomarker for electroconvulsive therapy outcome in depression: a multicenter machine learning analysis

**DOI:** 10.1101/2021.07.29.21261206

**Authors:** Willem B. Bruin, Leif Oltedal, Hauke Bartsch, Christopher C. Abbott, Miklos Argyelan, Tracy Barbour, Joan A. Camprodon, Samadrita Chowdhury, Randall Espinoza, Peter C. R. Mulders, Katherine L. Narr, Mardien L. Oudega, Didi Rhebergen, Freek ten Doesschate, Indira Tendolkar, Philip van Eijndhoven, Eric van Exel, Mike van Verseveld, Benjamin Wade, Jeroen van Waarde, Paul Zhutovsky, Annemiek Dols, Guido A. van Wingen

## Abstract

**Background:** Electroconvulsive therapy (ECT) is the most effective intervention for patients with treatment resistant depression. A clinical decision support tool could guide patient selection to improve the overall response rate and avoid ineffective treatments with adverse effects. Initial small-scale, mono-center studies indicate that both structural magnetic resonance imaging (sMRI) and functional MRI (fMRI) biomarkers may predict ECT outcome, but it is not known whether those results can generalize to data from other centers.

**Objective:** To develop and validate neuroimaging biomarkers for ECT outcome in a multi-center setting.

**Methods:** Multimodal data (i.e., clinical, sMRI and resting-state fMRI) was collected from seven centers of the Global ECT-MRI Research Collaboration (GEMRIC). We used data from 189 depressed patients to evaluated which data modalities or combinations thereof could provide the best predictions for treatment response (≥50% symptom reduction) or remission (HAM-D score ≤7) using a support vector machine classifier.

**Results:** Remission classification using a combination of gray matter volume and functional connectivity led to good performing models with average 0.82-0.83 area under the curve (AUC) when trained and tested on samples coming from the three largest centers, and remained acceptable when validated using leave-one-site-out cross-validation (0.70-0.73 AUC).

**Conclusions:** These results show that multimodal neuroimaging data is able to provide good prediction of remission with ECT for individual patients across different treatment centers, despite significant variability in clinical characteristics across centers. This suggests that these biomarkers are robust, indicating that future development of a clinical decision support tool applying these biomarkers may be feasible.

## Introduction

Electroconvulsive therapy (ECT) is currently the most effective intervention for patients with treatment resistant depression. Despite its high efficacy, ECT remains underutilized, as only 1-2% of patients with severe or persistent depression receive ECT (1). Although approximately 48% of treatment resistant patients recover with ECT, it is also associated with adverse cognitive effects and may be regarded as more invasive than other treatment options because the use of anesthesia is essential (2). Furthermore, ECT is relatively expensive and non-responsiveness can only be determined after multiple sessions. Information that better predicts treatment outcome would enable patient selection thereby further improving the overall response rate and avoiding ineffective treatment with adverse effects. A personalized recommendation about the expected benefit of ECT would be a valuable addition to the treating physician’s clinical judgement, and may increase its use in clinical practice.

Meta-analyses of studies that investigated predictors of ECT outcome have associated several clinical characteristics with beneficial ECT outcome, in particular no history of treatment resistance, older age and presence of psychotic symptoms (3, 4). However, the effect sizes are small, limiting their use to guide individual patient selection. Recent studies have started using neuroimaging data to predict ECT outcome at the individual level using machine learning analysis, which can construct multivariate prediction models using all the available data. Initial small-scale studies have shown that both structural magnetic resonance imaging (MRI) and functional MRI findings can be used to predict ECT outcome with approximately 80% accuracy (5, 6), which is considered sufficient for clinical use (7). These initial results have been confirmed by subsequent studies, and a recent meta-analysis showed an average prediction accuracy of 82% (8).

Despite these promising results, the existing studies have been limited by small sample sizes and mono-center settings. This reduces the possibility for models to generalize to new patients across centers. Although machine learning models typically perform better when trained on large samples from the same center, classification accuracy of larger multicenter studies tends to decrease, presumably due to increased clinical (e.g., adults vs. elderly) and technological (e.g., different MRI hardware and protocols) variability across centers (9). In order to develop robust and generalizable neuroimaging biomarkers for ECT outcome, we used data from the Global ECT-MRI Research Collaboration (GEMRIC) and validated classification performance in a multi-center setting (10). We used multimodal data (i.e., clinical, structural MRI (sMRI), and resting-state functional MRI (rs-fMRI)) and evaluated which data modalities or combinations thereof might provide the best predictions. As previous studies and clinical trials have used either treatment response (at least 50% symptom reduction) or remission (17-item Hamilton Depression Rating Scale (HAM-D) score of ≤7 after treatment) as outcome criterion, we assessed prediction accuracy for both criteria. Additionally, as most sites only contributed a small sample, we also evaluated model performance when only data from centers with ≥20 patients were used to provide classifiers with sufficient examples per center, which potentially could increase classification performance. Finally, we visualized the brain regions that were most informative to the classifications, in order to gain insight into the brain regions predictive of ECT outcome. To adhere to guidelines on reporting of diagnostic studies, we report our findings based on TRIPOD guidelines (11)

## Methods

### Participants

We performed a retrospective study using data from GEMRIC (v3.1, DOI:10.17605/OSF.IO/WD436), an international consortium that contains the largest multi-center database of neuroimaging scans on ECT (10). All contributing sites received ethics approval from their local ethics committee or institutional review board. In addition, the centralized mega-analysis was approved by the Regional Ethics Committee South-East in Norway (No. 2018/769). Analyses contained a selection of sMRI and rs-fMRI data from seven centers across Europe and North America, recorded from 189 clinically depressed patients according to ICD-10 (167 unipolar, 22 bipolar) who had received right unilateral or bilateral ECT (or both; **Supplementary Table 1**). Depressed patients were eligible for ECT treatment, typically after failure to respond to first-line treatments with conventional psychotherapy and antidepressant medications. The patients were included because of the availability of both high quality sMRI and rs-fMRI data. ECT stimulus parameters varied between different centers, including electrode placement. A description of center-specific ECT procedures and image acquisition is provided elsewhere (10). As GEMRIC consists of samples ranging from very small to relatively large (N=14, 14, 15, 18, 19, 29, 38, 42), we performed all analyses on the entire cohort or on centers with ≥20 patients available (three centers, N=109) in order to ensure classifiers were provided with sufficient data per center.

### Choice of primary measure

Treatment outcome was measured using the HAM-D or Montgomery-Åsberg Depression Rating Scale (MADRS) that was converted to HAM-D (**Supplementary Methods**), which are gold standard ratings for depression severity. Treatment response was defined as ≥50% HAM-D decrease compared to baseline and remission (minimal symptoms) as post-ECT HAM-D score ≤7.

### MRI data and preprocessing

MRI acquisition parameters are listed in **Supplementary Tables 2-3**. Structural T1-weighted scans were acquired using 1.5T and 3T scanners with a minimum resolution of 1.33mm^3^ and preprocessed using the CAT12 toolbox (v12.6; http://www.neuro.uni-jena.de/cat/) for voxel-based morphometry (VBM). Images were segmented into gray matter (GM), white matter (WM) and cerebrospinal fluid (CSF), normalized to MNI space using DARTEL registration, resampled to 1.5mm^3^ isotropic and spatially smoothed with an 8mm isotropic Gaussian kernel. GM data were masked at 0.2 to exclude WM.

150-266 rs-fMRI volumes were acquired with a TR of 1.7-3.0s, in-plane resolution of 2.4-3.75mm, and slice thickness of 3-5mm. Preprocessing was performed using ANTs (v2.2.0; https://github.com/ANTsX/ANTs) and FSL (v5.0.10; http://fsl.fmrib.ox.ac.uk/), including brain extraction, boundary-based co-registration, motion correction, spatial smoothing with a 5mm isotropic Gaussian kernel, and normalization to a 2mm MNI template. Denoising was performed using ICA-AROMA, and depending on the type of analysis, high-pass (f>0.01) or bandpass filtering (0.009<f<0.08) was applied together with WM and CSF nuisance regression (12). Denoised rs-fMRI data were resampled to 4mm isotropic.

Only subjects that passed quality control for both rs-fMRI and sMRI were included for analysis, leading to a final sample of 189 patients (**Supplementary Figure 1** for a flowchart). Details on MRI preprocessing, quality control and machine learning are provided in the **Supplementary Methods**.

### Feature extraction

We extracted commonly used MRI features from the preprocessed data. For sMRI, we used voxel-wise modulated GM maps (VBM) and 142 cortical and subcortical parcellations using the Neuromorphometrics atlas (NMM; provided by Neuromorphometrics, Inc). For rs-fMRI, we used a high-dimensional resting-state networks template from the UK BioBank dataset to extract 100 independent spatial components that were derived using group independent component analysis (ICA) (13). 45 components reflecting non-neural signals and three components mainly located in cerebellar regions with insufficient EPI coverage were discarded, resulting in 52 components for analysis. Group-information guided ICA was used to derive subject-specific time-series and spatial maps for each of the 52 signal components using the high-pass filtered preprocessed data (14). Time-series were used to calculate individual functional connectivity (FC) matrices that described pairwise connectivity between signal components with Pearson correlations (ICA FC). Additionally, we used an atlas-based approach from Power *et al*., and extracted time-series from 264 functional areas to compute FC matrices (Power FC) using the band-pass filtered preprocessed data (15). Correlations were converted to z-scores with Fisher r-to-z transformation before performing the classification.

### Machine learning

Machine learning classifications were performed using linear support vector machine classifiers (SVM; LIBSVM for Python (https://www.csie.ntu.edu.tw/~cjlin/libsvm/) implemented in scikit-learn (v0.19.1; https://scikit-learn.org/)) and validated using stratified shuffle-split cross-validation (CV) with 100 iterations. At each iteration, stratified-splits were made by preserving the proportion of responders/remitters and non-responders/remitters from each center to obtain maximally homogeneous train-test splits in which 80% data was used for classifier training and 20% for testing. This CV procedure is further referred to as ‘internal validation’. In addition, we performed leave-one-site-out (LOSO) CV, in which all but one center was used to train the SVM while the remaining center was used to assess model performance (further referred to as ‘external validation’). This procedure was repeated so that each center was used once for testing. LOSO reduces the risk of overfitting to data from a single center but may result in large between-sample heterogeneity of training and test sets, which could result in lower classification performance compared to internal validation. Hyper-parameters for the SVM were optimized using grid-search via nested cross-validation with 10 iterations. We assessed classification performance using different sets of MRI features (VBM, NMM, ICA-DR FC, Power FC, and ICA spatial components), as well as using clinical data only (i.e., age, sex and pre-ECT HAMD scores) for baseline classification. Clinical data were always included for each classification. The primary performance metric was area under the receiver operator characteristic curve (AUC) and reported metrics were averaged across CV iterations. Balanced accuracy, sensitivity, specificity, positive predictive value (PPV) and negative predictive value (NPV) are reported in **Supplementary Tables 6-13**.

Statistical significance of classification performance was assessed using label permutation-testing with 1000 iterations (16). Obtained p-values were corrected for multiple comparisons using False Discovery Rate (FDR; two-stage (non-negative); alpha=0.05). 95% confidence intervals (CI) for AUC were computed using the modified Wald-method (17). To reduce the computational burden, only spatial ICA classifications that resulted in AUC>0.75 were tested for significance. Finally, we assessed classification performance for multi-modal classifications combining anatomical and functional features: regional neuromorphometrics GM volumes with either ICA or Power-atlas based FC, and voxel-wise GM with either ICA or Power-atlas based FC.

### Anatomical localization

To investigate which regions contributed most to the voxel-wise classification, we employed a method to estimate p-values for the weights of the SVM (18). A statistic was computed incorporating the SVM weight component value and the size of the margin, and an analytical approximation to the null-distribution obtained through permutation testing was used to calculate p-values.

## Results

### Demographic data

Demographic data is presented in Table 1. Of the 189 included patients, 113 patients were ECT responders and 76 non-responders, and 76 were remitters and 113 non-remitters. As expected, patients with a favorable outcome were older, and higher symptom severity at baseline was associated with ECT response but not remission. No significant differences in sex, initial electrode placement and total number of ECT-sessions were observed.

**Table 1.** Demographics of patients included in data analysis, with subject demographics and comparisons between ECT responders and non-responders, and between remitters and non-remitters. Abbreviations: m: male; f: female; RUL: right unilateral ECT initially, BL: bilateral ECT initially; HAM-D: Hamilton Rating scale for depression; UP: unipolar depression; BP: bipolar depression; n.a.: not available. Asterisks depict significance using independent t-test or χ ^²^ test.

|  | Total sample (n=189) |  | Responders (n=113) |  | Non-Responders (n=76) |  | p | Remitters (n=76) |  | Non-Remitters (n=113) |  | p |
| --- | --- | --- | --- | --- | --- | --- | --- | --- | --- | --- | --- | --- |
|  | mean | std | mean | std | mean | std |  | mean | std | mean | std |  |
| Age | 51.7 | 14.5 | 54.400 | 13.7 | 47.6 | 16.9 | 0.0045* | 56.3 | 14.2 | 48.6 | 15.5 | 0.0006* |
| Sex (m/f) | 83/106 | n.a. | 52/61 | n.a. | 31/45 | n.a. | 0.5705 | 32/44 | n.a. | 51/62 | n.a. | 0.7935 |
| Laterality (RUL/BL; n=188) | 148/40 | n.a. | 87/25 | n.a. | 61/15 | n.a. | 0.8 | 60/15 | n.a. | 88/25 | n.a. | 0.86 |
| HAM-D pre-treatment | 25 | 7.7 | 26.300 | 7.3 | 23 | 7.7 | 0.0031* | 25.8 | 8.2 | 24.5 | 7.2 | 0.26 |
| HAM-D post-treatment | 11 | 8.3 | 5.700 | 4.2 | 18.9 | 6.3 | 5.82E-31* | 3.3 | 2.3 | 16.2 | 6.6 | 3.73E-42* |
| HAM-D change | 14 | 10.7 | 20.600 | 7.7 | 4.1 | 5.8 | 6.31E-39* | 22.5 | 8.3 | 8.2 | 7.8 | 3.77E-23* |
| Diagnosis (UP/BP) | 167/22 | n.a. | 99/14 | n.a. | 68/8 | n.a. | 0.8726 | 67/9 | n.a. | 100/13 | n.a. | 0.8726 |
| Total ECT sessions (n=186) | 13.4 | 6.2 | 13.000 | 6.4 | 14 | 5.8 | 0.2665 | 12.9 | 6.7 | 13.8 | 5.8 | 0.3474 |

We assessed differences in sample demographics and clinical characteristics between different centers regardless of ECT outcome using one-way analysis of variance (ANOVA) and χ^2^. Age (F(7,181)=14.08, p<0.001), pre-treatment HAMD scores (F(7,181)=7.40, p<0.001), post-treatment HAMD scores (F(7,181)=5.24, p<0.001), HAM-D change (F(7,181)=8.65, p<0.001), number of ECT sessions (F(7,178)=10.78, p= p<0.001), depression type (*X*^*2*^(7, N=189)=19.10, p=0.008) and initial electrode placement laterality (*X*^*2*^(7, N=189)=109.8, p<0.001) differed significantly between centers. In contrast, sex did not differ between centers (*X*^*2*^(7, N=189)=3.84, p=0.80). Demographic data for the three largest centers (with N≥20) used for additional analyses are described in **Supplementary Tables 4-5**. Differences in sample demographics and clinical characteristics between the three largest centers were similar to those seen in the entire sample. These findings highlight that there is considerable clinical heterogeneity between centers.

### Remission prediction

#### All centers

##### Unimodal neuroimaging

Performance for remission classification was evaluated with internal stratified cross-validation (stratified-shuffle splits) across centers using all samples (N=189). Remission classification performance was poor with AUCs (averaged across 100 CV-iterations) ranging between 0.58-0.67 for clinical data only (age, sex and pre-ECT HAM-D scores) and VBM, NMM, ICA-FC and Power-FC feature sets that also included the clinical data in all analyses (Figure 1A). Nonetheless, these AUCs except for NMM were statistically significant following permutation testing with multiple comparison correction. Classification using external (LOSO) cross-validation hardly exceeded chance-level, with AUCs (averaged across sites) ranging between 0.51-0.58 and none were statistically significant (Figure 1B). Classification using ICA networks did not exceed AUC>0.75 for either internal or external validation. Notably, one ICA component centered around the anterior temporal lobes that included the amygdala and hippocampus resulted in 0.70 AUC but did not obtain statistical significance following with multiple comparison correction (p_FDRcorrected_=0.2078; p_uncorrected_=0.0019) (Figure 2A). More comprehensive classification results, including balanced accuracy, sensitivity, specificity, PPV and NPV, p-values for AUC statistical significance and 95% CI’s are provided in **Supplementary Table 6**.

**Figure 1.**
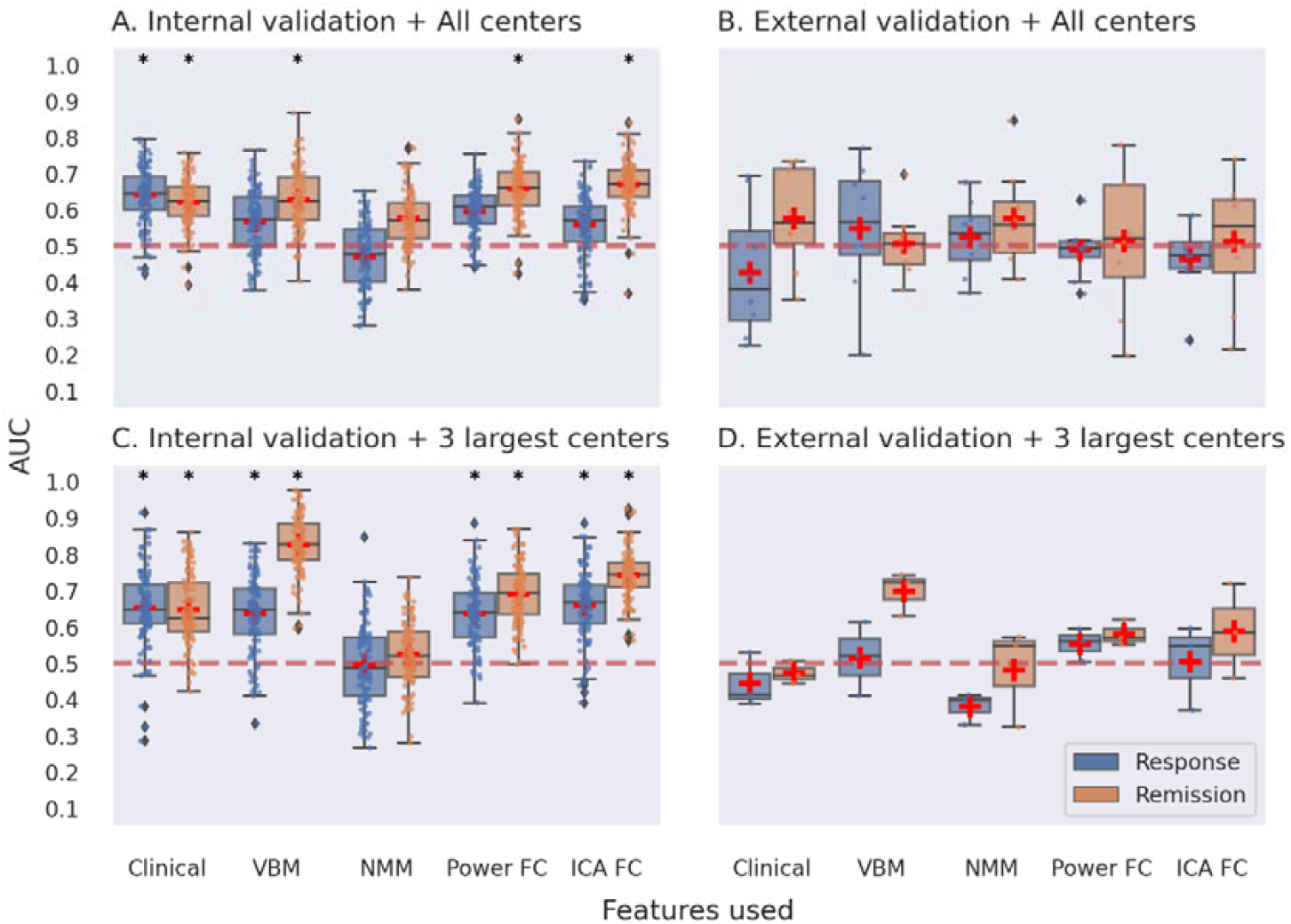
Multi-center predictions for ECT treatment response and remission using unimodal MR data modalities. Panel A depicts classification performance using data from all centers and different MR modalities with internal validation (AUC is averaged over 100 stratified cross-validation splits). Panel B shows classification performance using data from all centers with external validation (leave-one-site-out cross-validation, scores are averaged across different center left out for model testing). Panel C depicts classification performance using data from the three largest centers with internal validation. Panel D shows classification performance using data from the three largest centers with external validation. VBM = voxel-based morphometry; NMM = Neuromorphometrics atlas; FC = functional connectivity; ICA = group information guided independent component analysis. Red dashed line depicts chance level performance (0.5 AUC). Asterisks indicate significant difference from chance level after permutation testing with false discovery rate correction for multiple comparisons (p<0.05, corrected).

**Figure 2.**
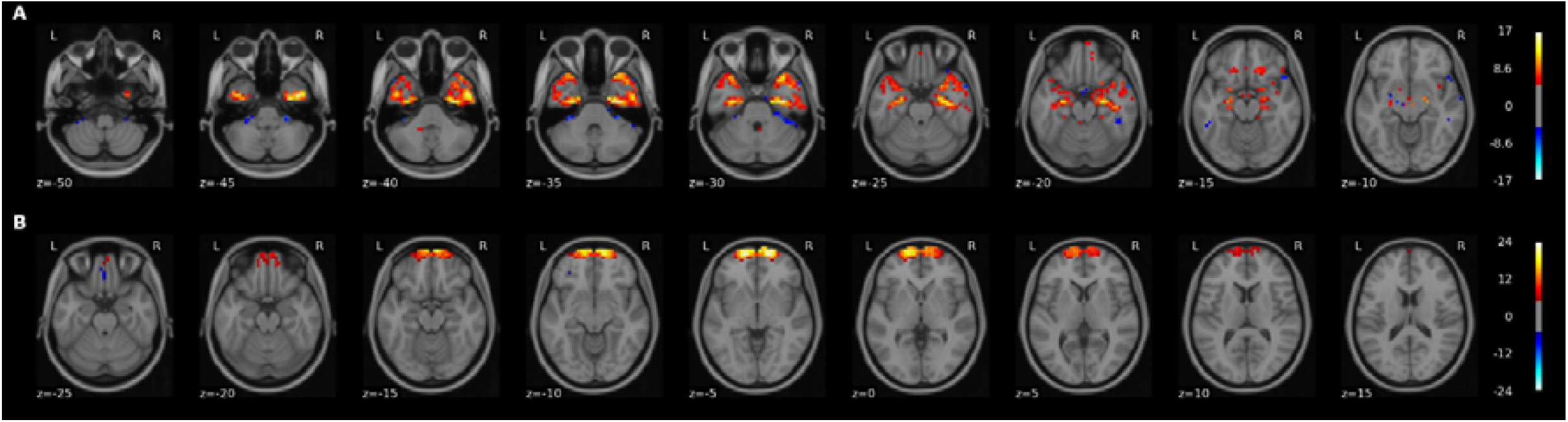
Visual representation of the two UK BioBank group ICA spatial components that led to AUC>0.75 for either response or remission classification. Top panel A depicts a network (#42) centered around the temporal lobes. The second panel B shows a network (#52) located in frontopolar cortex. Images are thresholded at Z≥5 and overlaid on a standard 2mm MNI template. The Figure was made with the nilearn package (http://nilearn.github.io).

##### Multimodal neuroimaging

Classification using a combination of anatomical MRI, functional MRI and clinical data led to a maximum of 0.68 AUC using internal validation which was statistically significant, and a maximum of 0.64 AUC for external validation which did not obtain significance (Figure 3A, 3B; **Supplementary Table 7**).

**Figure 3.**
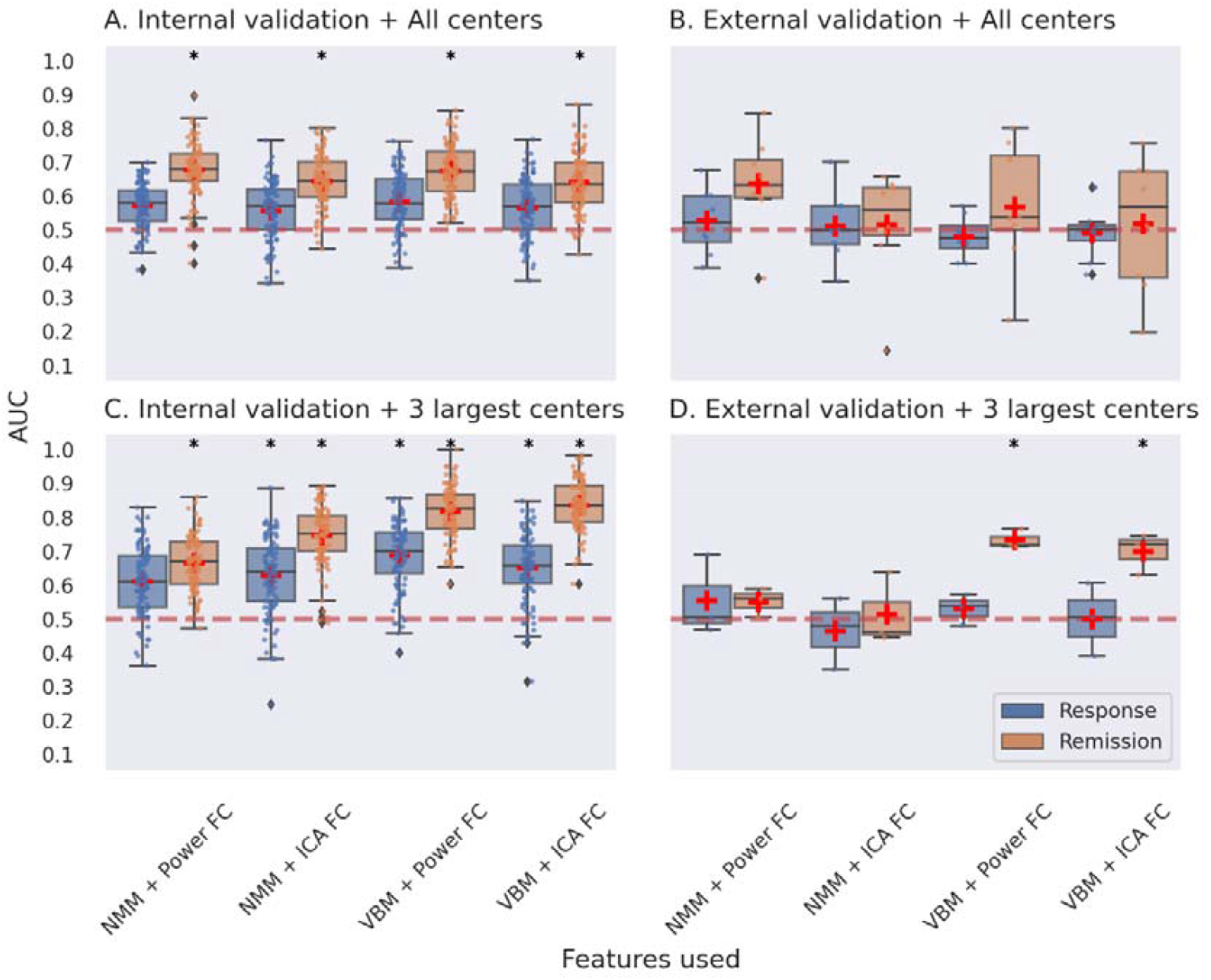
Multimodal multi-center predictions for ECT response and remission. Panel A depicts classification performance using data from all centers and different combinations of features with internal validation (AUC is averaged over 100 stratified cross-validation splits). Panel B shows classification performance using data from all centers and different combinations of features with external validation. Panel C depicts classification performance using data from the three largest centers with internal validation. Panel D shows classification performance using data from the three largest centers with external validation. VBM = voxel-based morphometry; NMM = Neuromorphometrics atlas; FC = functional connectivity; ICA = group information guided independent component analysis. Red dashed line depicts chance level performance (0.5 AUC). Asterisks indicate significant difference from chance level after permutation testing with false discovery rate correction for multiple comparisons (p < 0.05, corrected).

#### Three largest centers

##### Unimodal neuroimaging

We next assessed prediction performance only using data from three centers with N≥20 (N=109) to provide the machine learning classifier with sufficient samples per center. Classification performance with internal validation ranged between 0.52-0.83 AUC across different features used, and 0.65 AUC was obtained for classifications using clinical variables only (Figure 1C). All these AUCs, except for NMM, showed statistical significance. Notably, the highest performance was achieved using voxel-wise GM data with 0.83 AUC. Two out of 52 ICA networks resulted in AUC>0.75 (Figure 2). One component centered around the temporal lobes resulted in 0.75 AUC, and a frontopolar network resulted in 0.80 AUC, but neither obtained statistical significance after multiple comparison correction. Classifications performed with external validation ranged between 0.47-0.70 AUC (Figure 1D). The performance obtained with voxel-wise GM data using internal validation was reduced from 0.83 AUC to 0.70 AUC with external validation, and failed to obtain statistical significance following permutation testing with multiple comparison correction (p_FDRcorrected_=0.0899; p_uncorrected_0.0089). None of the ICA networks resulted in AUC>0.75 with external validation (**Supplementary Table 8**).

##### Multimodal neuroimaging

Classification combining voxel-wise GM with ICA-based FC led to the best performing model, with 0.83 AUC using internal validation and 0.70 AUC using external validation. Classifications for voxel-wise GM with the Power-atlas FC led to similar performances of 0.82 AUC for internal validation and 0.73 AUC for external validation. All of the aforementioned AUCs were statistically significant for both internal and external validation (Figure 3C, 3D). Classification performance for regional NMM with ICA-based FC resulted in 0.75 AUC with internal validation and 0.51 AUC for external validation. Classifications for regional NMM with Power-atlas FC led to 0.67 AUC using internal validation and 0.55 AUC for external validation. AUCs obtained for classifications using regional neuromorphometrics and FC were statistically significant for internal validation but not for external validation (**Supplementary Tables 9**).

### Response prediction

#### All centers

##### Unimodal neuroimaging

Overall response classification using data from all sites was poor, with AUCs ranging between 0.47-0.62 for different MRI features used (Figure 1A). Classification using clinical variables only resulted in a comparable AUC of 0.64. All classification performances, except for those using clinical variables only, were not significant. Classification performances evaluated with external validation did not exceed chance-level with AUCs ranging between 0.43-0.56 (Figure 1B). A full overview of these results can be found in **Supplementary Table 10**.

##### Multimodal neuroimaging

Classification using a combination of anatomical and functional MRI features led to a maximum of 0.58 AUC using internal validation, and 0.53 AUC for external validation, which were not significant (Figure 3A, 3B; **Supplementary Table 11**).

#### Three largest centers

##### Unimodal neuroimaging

Classification with internal validation on the three largest centers remained poor with AUCs ranging between 0.5-0.69 across MRI features used, and 0.65 AUC for classifications using clinical data only (Figure 1C). Notably, classifications using ICA-FC and VBM data performed best, with 0.66 and 0.64 AUC respectively, and both were found to be statistically significant. Classifications evaluated using external validation ranged between 0.38-0.55 and none were statistically significant (Figure 1D; **Supplementary Table 12**).

##### Multimodal neuroimaging

Classification performances with internal validation were higher with AUCs ranging between 0.61-0.69, but remained poor using external validation with a maximum AUC of 0.55 **(Supplementary Tables 13)**.

#### Learning curves

To evaluate the relation between training sample size and classification performance, we examined learning curves for the best performing models (i.e. remission classification using data from the three largest centers) by using different proportions of training data. Classification accuracy reached 0.83-0.84 AUC for unimodal (voxel-wise GM) and multimodal (voxel-wise GM and ICA-based FC) classifiers, with averaged AUC>0.75 for classifications using 50% of data for training (N=55) and AUC>0.8 for 60% of data used for training (N=66). See **Supplementary Figure 2** for full learning curves. Both learning curves did not appear saturated, suggesting that model performance could still increase when using larger training samples.

#### Anatomical localization

We investigated which brain regions contributed most to treatment classification for voxel-wise GM data. We only focus on our best performing unimodal model, which for remission classification resulted in 0.83 AUC using data from the three largest samples. P-values were plotted for GM weights only as we were interested in brain regions rather than the influence of covariates. As shown in Figure 4, regions located in dorsomedial prefrontal (dmPFC), precuneus and thalamus exhibited high contribution to the classification task. The sign of weights within thalamus was mostly negative, implying a high chance for non-remission classification, whereas signs of weights within dmPFC and precuneus were mostly positive, implying a high chance for remission classification. Note that these results reflected the contribution of these brain regions to the multivariate pattern used by the SVM classifier.

**Figure 4.**
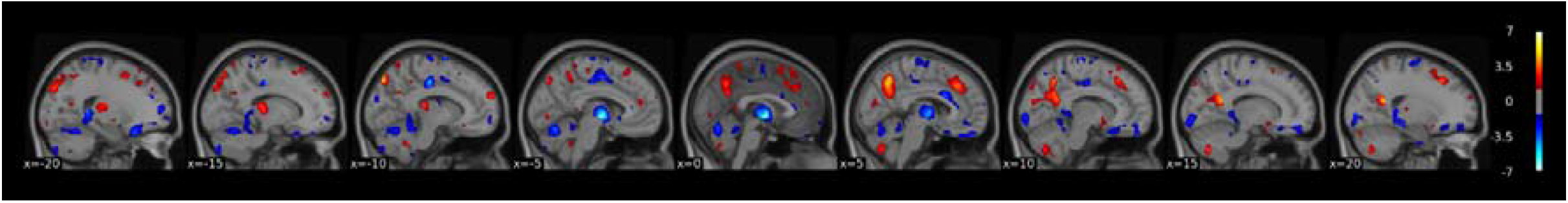
Thresholded −log(p) value maps characterizing the regions important for the treatment remission classification using voxel-wise GM data of the three largest centers (thresholded at p<0.05 uncorrected). Hot colors indicate positive weights and cold colors indicate negative weights of the SVM. The Figure was made with the nilearn package (http://nilearn.github.io).

## Discussion

The presented results show that neuroimaging data can provide a good prediction of ECT remission for individual patients across different centers. In line with recent meta-analyses, older age and higher depression severity at baseline were associated with better ECT outcome (3). However, our classification results show that this information is not sufficient for making individual predictions, highlighting the relevance of obtaining neuroimaging data for accurate predictions. Remission classification using a combination of voxel-wise GM and clinical data with either ICA-based FC or Power-atlas based FC led to good performing models when trained and tested on samples coming from each center (internal validation AUC>0.8), and remained acceptable when validated on completely new data from other centers (external validation AUC>0.7) (19). These results indicate that multimodal neuroimaging data may provide a robust biomarker that could be used to guide clinical decision-making. By providing patients and clinicians a patient-specific prognosis, this could ultimately increase the success rate of ECT, avoid ineffective treatments and accompanying adverse effects, and increase the use of the most effective antidepressive treatment available.

Previous monocenter studies using neuroimaging data to predict ECT outcome with either structural or functional MRI were able to obtain up to 0.83 AUC (8). Here we achieved similar classification performance in a multicenter setting. Using data from different samples involves many additional sources of technological (e.g., different MR hardware and scanner protocols) and clinical (e.g. different ECT protocols, patient cohort and recruitment procedures) variability (9). These additional sources of variability may decrease prediction accuracy of MRI measurements for ECT outcome (9). Conversely, a multicenter study avoids cohort-specific solutions and so helps test generalizability of the results across different samples, increasing the likelihood that features identified as discriminatory between remitters and non-remitters reflect generic properties related to treatment outcome across datasets. Our results showed that generalizability to new samples came at the cost of lower accuracy, as best performing classifications with internal validation (AUC≈0.83) outperformed those using external validation (AUC≈0.73). Additionally, we found that using a subsample of the data containing three centers with N≥20 each (N=109) led to better model performance compared to using all seven centers (N=189). This improvement could not be attributed solely to reduced clinical heterogeneity, as differences in sample demographics and clinical characteristics between the three largest centers were found to be similar to those seen in the entire sample (**Supplementary Tables 1-2**). We therefore hypothesize that the exclusion of smaller centers ensured that the model had sufficient examples per center for training.

Brain regions that contributed most to remission classification using structural MRI data included dmPFC, precuneus and thalamus. The thalamus is a hub connecting all cortico-cortical circuits with links to hippocampus and medial PFC. It is a central hub in the affective network and plays an important role in emotion dysregulation (20). There is evidence of decreased thalamic volume and hyperactivity during rest in depression (21, 22), and thalamic volume and function have been associated with ECT efficacy (23, 24). Preliminary evidence links changes in precuneus network connectivity and structure with ECT treatment outcome (25, 26). A previous ECT outcome prediction study using structural MRI also implicated the precuneus (27), and studies using rs-fMRI have reported thalamic connectivity as an important predictor (23, 28).

Our results also indicated an important role for remission classification using FC within resting-state components centered around the anterior temporal lobes and frontopolar cortex. The frontopolar cortex (Brodmann Area 10) plays an important role in integrating cognitive, social, and emotional processes. Its medial parts are mostly associated with affective processing such as emotional and social cognition, and it lateral parts with working memory and perception. (29, 30). Previous studies have reported that depression is associated to reduced medial frontal pole volume (31) and that decreased frontal pole volume and FC following ECT was related to therapeutic efficacy (32). The medial temporal lobes have been consistently implicated in ECT neuroimaging research and include the hippocampus and amygdala, which have shown to undergo structural changes in volume, functional connectivity and perfusion following ECT treatment (5, 24, 26, 33–35). The temporal lobes also show the highest magnitude of electrical current in RUL stimulation (36), and increased electrical field strength has been associated with increased right hippocampal neuroplasticity and improved antidepressant outcomes (37). Smaller hippocampal volumes, and to a lesser extend the amygdala, are apparent in MDD patients and support the current hypothesis that mood disorders consist of dysfunction in neural circuits important for processing and integrating emotional and cognitive events (38, 39). Previous classification studies have also highlighted anterior lateral temporal lobe volume, hippocampal and amygdala gray matter (27, 40, 41), and temporal cortex functional connectivity (42) as important predictors for ECT outcome. Altogether, these results provide evidence for the importance of thalamic and precuneus structure and fronto-temporal FC for both depression and ECT-related clinical response.

Notably, the identification of brain regions contributing most to the classification resulted from a multivariate analysis, and the localization of these regions should therefore be interpreted with caution as these regions may not only be related to treatment outcome but also contribute to denoising during the classification process (43). Several limitations have to be taken into account when interpreting our findings. We used a retrospectively pooled sample from existing data across the world, without harmonized protocols for scanning, inclusion criteria or demographic and clinical characteristics. Not surprisingly, we found significant differences in sample demographics and clinical characteristics between the different data collection centers. These sources of heterogeneity may limit classification performance but also provide an opportunity for model development using independent data sets and the discovery of generalizable biomarkers that are reproducible across centers. However, classification performance might be improved by using standardized acquisition parameters for possible future clinical utility. Additionally, our findings show that the prediction of treatment response was poor, while prediction of remission was good. This indicates that ECT outcome prediction is limited to remission and that the remission group can be best differentiated from the other patients. Remission may provide a better outcome criterion than response and has become the gold standard for depression treatment, because patients who do not remit have a poorer prognosis and greater chance of relapse and recurrence than those who do. Remission is also associated with a lower full symptomatic recurrence rate compared with achieving treatment response (3, 44). Furthermore, while unimodal and multimodal models performed comparable for remission classification using data from the largest centers with internal validation, only the multimodal classifications remained acceptable with external validation on different centers. We speculate that multimodal data may increase the probability that either the structural or functional MRI data overlaps across centers.

Taken together, this study suggests that ECT remission can be accurately predicted using MRI data in a large, ecologically valid, multi-center sample of patients receiving ECT, indicating that future development of a clinical decision support tool might be feasible. MRI could easily be incorporated during decision making, as ECT is typically provided in a hospital setting. And as MRI is inexpensive compared to ECT, the additional costs are expected to outweigh the costs of unsuccessful treatments.

## Supporting information

TRIPOD checklist

## Data Availability

We used data from GEMRIC, an international consortium that contains multi-center database of neuroimaging on ECT. GEMRIC data will only be shared with the collaborators in GEMRIC, and data will be stored on a secure server at the University of Bergen (UoB), Norway. Collaborators can upload, access and work with the data through a secure connection, provided by UoB. It is not allowed to download the raw data from the server. All participating sites must have ethical approval for their projects and for data sharing. More info found here: https://mmiv.no/how-to-join-gemric/

## Acknowledgements

We would like to thank the logistic and academic support of the entire GEMRIC consortium. The full overview of the GEMRIC board members can be found here: https://mmiv.no/gemric/. In addition, we would like to acknowledge Louise Emsell for contributing data that was considered but not used for the final analysis. This work was supported by the Netherlands Organization for Scientific Research (NWO/ZonMW Vidi 917.15.318, Dr. van Wingen), Western Norway Regional Health Authority (Grant No. 91223, Dr. Oltedal), NARSAD Young Investigator Grant (No. 27786 to BW) and a K99 Pathway to Independence Award (Grant No. MH119314 to BW), the National Institute of Mental Health (Grant No. MH092301 and MH110008 for Dr. Narr and Dr. Espinoza; MH111826 and MH125126 for Dr. Abbott; MH119616 for Dr. Argyelan and R01MH112737 for Dr. Camprodon).

## Contributors

WBB, GAvW & AD conceived the research question, contributed to the design and wrote the first draft of the manuscript. WBB performed data analysis and developed MRI processing pipelines. GAvW and PZ made substantial contributions to data analysis. LO coordinated the GEMRIC consortium. WBB, LO, HB, CCA, MA, TB, JAC, SC, RE, PCRM, KLN, MLO, DR, FtD, IT, PvE, EvE, MvV, JvW, AD & GavW contributed to data acquisition. WBB, PZ, LO, HB, CCA, MA, TB, JAC, SC, RE, PCRM, KLN, MLO, DR, FtD, IT, PvE, EvE, MvV, BW, JvW, AD & GavW substantially revised the manuscript. All authors critically revised the manuscript for important intellectual content, approved the final draft and had final responsibility for the decision to submit for publication.

## Data sharing statement

Individual participant data cannot be made available publicly because there is no consent or ethical approval for this and the data cannot be anonymized. The data are stored on a secure centralized server at the University of Bergen, Norway. Participating GEMRIC sites have access to the raw data according to specific data policy and safety rules of the consortium, and in accord with the approval from the ethical committee. The GEMRIC consortium welcomes new members who are interested in the neuroimaging research of ECT. For more about the application process please visit https://mmiv.no/how-to-join-gemric/ or write to Leif Oltedal. General information about the consortium can be found on the following website: https://mmiv.no/gemric/.

