## Supplementary File for "Development and validation of a multimodal neuroimaging biomarker for electroconvulsive therapy outcome in depression: a multicenter machine learning analysis"

### **S1 Supplementary Methods**

### **HAM-D conversion**

For centers collecting only MADRS scores, a validated equation was used to convert them to the 17-item Hamilton Depression Rating score by Heo *et al.,* 2007 (1):

$$HAM-D = -1.58 + (0.86 * MADRS)$$

**MRI data and preprocessing**

Structural T1-weighted (T1w) MRI scans with a minimum resolution of 1.33 mm^3^ were acquired before and after ECT using either a 1.5T or 3T scanner (full image acquisition parameters are listed in **Supplementary Tables 2-3**). Structural MRI preprocessing was performed using the CAT12 toolbox (v12.6; <http://www.neuro.uni-jena.de/cat>) for voxel-based morphometry (VBM) and SPM12 (v7487; <http://www.fil.ion.ucl.ac.uk/spm>) in MATLAB (R2019A; <http://www.mathworks.com>). Images were first corrected for scanner-specific gradient nonlinearity and reoriented by centering the image on the anterior commissure (2). Preprocessing consisted of tissue-segmentation into gray matter (GM), white matter (WM) and cerebrospinal fluid (CSF) volumes following normalization to Montreal Neurological Institute (MNI) space using a DARTEL registration with the CAT12 template derived from 555 HC subjects of the IXI-database (<http://www.brain-development.org>) (3). Normalized images were then modulated with the determinant of Jacobian matrices of the deformations, computed during the nonlinear registration, to compensate for effects of volume changes caused by affine registration and non-linear warping, so that resulting images reflected regional tissue volumes before spatial normalization (4). Normalized GM volumes were smoothed with an 8 mm full-width-at-half-maximum Gaussian kernel to suppress noise and effects due to variation in (gyral) anatomy caused by inter-subject averaging. A GM mask was created by thresholding the individual GM images at 0.2 thereby ensuring that a voxel was only included if at least 51% of patients included the same voxel in their individual mask. Finally, scans were manually checked for correct orientation and homogeneity tests were used to detect outliers with artefacts or poor quality using Mahalanobis distance boxplots that incorporated CAT12 segmentation quality reports and total intracranial volume (TIV) and age as nuisance variables.

Additionally, 150-265 volumes of rs-fMRI were acquired with a TR of 1.7-3.0 s, in-plane resolution of 2.4-3.75 mm, and slice thickness of 3-5 mm. Anatomical brain extraction was applied using ANTs (v2.2.0; using the OASIS template (5)) to remove non-brain tissue. Next, FMRI Software Library (FSL; v5.0.10) FEAT was run for boundary-based registration (BBR) co-registration using the brain extracted T1w (6). The first two volumes of the functional scan were discarded, motion correction was applied with respect to the middle volume (MCFLIRT) using six parameters for rigid body head motion transformations and spatial smoothing was applied on each volume separately using a Gaussian kernel of 5 mm full-width-at-half-maximum to reduce noise. Volumes were then normalized by a single scaling factor (grand mean scaling) so that each volume was scaled by the same amount. ANTs registration was used to compute nonlinear transformations from T1w to a 2 mm standard MNI template. Afterwards ICA-AROMA was applied to the preprocessed rs-fMRI data to remove additional motion sources. The estimated noise components identified by ICA-AROMA were then also used to denoise the cosine regressors used for high-pass (f>0.01) or bandpass filtering (0.009<f<0.08)(7). Additionally, mean WM and CSF time-series were computed as additional nuisance variables and similarly denoised by the noise components of ICA-AROMA. Both the denoised cosines and denoised WM and CSF nuisance variables were regressed out from the rs-fMRI data after ICA-AROMA. Finally, the preprocessed, denoised and band- or highpass filtered fMRI data were normalized to MNI standard space using the BBR co-registration and nonlinear transformations in one single step using *antsApplyTransforms* with Lanczos interpolation. The final preprocessed images were resampled to 4 mm isotropic to limit the number of voxels and speed up computations. The general level of motion in the rs-fMRI scans was assessed with relative frame-wise displacement (FD) estimates (6, 8). High-motion subjects were excluded based on the following three criteria: if any rotation/translation parameters exceeded 4 mm/degrees, if average FD exceeded 0.3 mm, or if subjects had less than 4 minutes of motion unaffected data (total scan duration of volumes with FD<0.25mm). Additionally, rs-fMRI and sMRI scans were visually inspected for quality control of co-registration, normalization, and EPI signal-to-noise and field-of-view to assess possible dropout and other artifacts.

Only data from subjects that passed quality control for both rs-fMRI and sMRI scans were included for analysis, leading to a final sample of 189 patients (see **Supplementary Figure 1** for a flowchart describing quality control).

**Feature extraction**

We extracted commonly used MRI features from the preprocessed data: voxel-wise GM maps and atlas-based regional GM volumes, and independent component analysis (ICA) and atlas-based functional connectivity (FC) were extracted from sMRI and rs-fMRI data, respectively. For sMRI we used voxel-wise modulated GM maps obtained from VBM analysis. Additionally, we used volumetric parcellations describing GM volume of 142 cortical and subcortical regions using the Neuromorphometrics atlas (NMM; from Neuromorphometrics, Inc.) provided by CAT12. Regional GM volume was extracted using a projection‐based thickness (PBT) approach that resembles FreeSurfer (v6) estimations, with similar intra-method repeatability and inter-method reproducibility (9). For rs-fMRI feature extraction we used an existing high-dimensional resting-state networks template obtained from the UK BioBank dataset derived using group independent component analysis (ICA) on approximately 4000-4500 subjects to extract 100 resting-state components (10). From these 100 components, 55 components were labeled as non-artifactual (i.e. components reflecting non-neural signals such as motion, WM and CSF). Out of the remaining 55 signal components, three components mainly located in cerebellar regions outside the group mask were discarded, resulting in 52 components considered for classification. Finally, group information guided (GIG-) ICA was used to derive subject-specific time-series and spatial maps for each of the 52 signal components (11). Time-series were used to calculate individual functional connectivity (FC) matrices that described pairwise connectivity between the 53 components with Pearson correlations. Additionally, we used an atlas-based FC approach using the Power coordinates describing 264 putative functional areas (12). Here, 4 mm spheres were extracted from the preprocessed data with bandpass filtering to derive averaged time-series for each of the 264 coordinates and then used to compute FC matrices. The upper triangular portion of the FC matrices were used as features, and its corresponding correlations were converted to z-scores by applying Fisher r-to-z transformation before entering classification. The total number of features used was: 406929 for voxel-wise VBM maps, 142 for NMM parcellations, 37401 for Power-based FC, 1378 for ICA-based FC, and 26629 for each of the 53 ICA spatial components identified as signal.

**Neuromorphometrics labels**

The neuromorphometrics atlas data were provided for use in the MICCAI 2012 Grand Challenge and Workshop on Multi-Atlas Labeling (B. Landman, S. Warfield, MICCAI 2012 workshop on multi-atlas labeling, in: MICCAI Grand Challenge and Workshop on Multi-Atlas Labeling, CreateSpace Independent Publishing Platform, Nice, France, 2012; provided by Neuromorphometrics, Inc.: [http:/neuromorphometrics.com/](http://www.neuromorphometrics.com/)). The atlas included 142 cortical, subcortical and ventricular regions. A full overview of the ROI labels and abbreviations can be found [here](https://www.jiscmail.ac.uk/cgi-bin/webadmin?A3=ind1806&L=UKB-NEUROIMAGING&E=base64&P=37625&B=--000000000000c942ed056e83bba8&T=application%2Fvnd.openxmlformats-officedocument.spreadsheetml.sheet;%20name=%22atlases_ROIs_list.xlsx%22&N=atlases_ROIs_list.xlsx&attachment=q&XSS=3).

**Machine learning classification**

Machine learning classifications were performed using a linear Support Vector Machine (SVM) classifier (LIBSVM (13) for Python, [https://www.csie.ntu.edu.tw/~cjlin/libsvm/](https://www.csie.ntu.edu.tw/~cjlin/libsvm/))), implemented in scikit-learn, and stratified shuffle split cross-validation (CV) with 100 iterations. At each iteration, stratified-splits were made by preserving the proportion of responders and non-responders (or remitters and non-remitters) from each center to obtain maximally homogeneous train-test splits in which 80% data was used for classifier training and 20% for testing. This CV procedure was further referred to as ‘internal validation’. In addition, we addressed leave-one-site-out (LOSO) cross-validation (CV), in which all but one center were used to train the models while the left out center was used to assess model performance (further referred to as ‘external validation’). This procedure was then repeated so that each center was used once as a test set. LOSO reduces the risk of overfitting data from a single center, and might result in large between-sample heterogeneity of training and test sets, which could result in lower classification performance compared to internal validation (14). Multisite classifications with internal and external validation enabled us to measure their respective impact on prediction.

Hyper-parameters for the linear SVM were optimized using nested CV: a grid-search was performed across different values of C (0.001, 0.01, 0.01, 01, 1, 10, 100) using 10 inner stratified shuffle splits. SVM class weights for C were set to “balanced” mode to automatically adjust weights inversely proportional to class frequencies in the input data. Decision function threshold optimization was performed by finding the decision threshold value that resulted in the largest Youden index, by using decision values obtained from another nested CV with 5 stratified shuffle splits. We assessed classification performance using different sets of extracted MRI features, as well as baseline classification using clinical data only (i.e. age, sex and pre-ECT HAMD scores). Baseline clinical data were always included for classification using MRI features by concatenating individual feature vectors. For feature scaling, age was divided by 100 and pre-ECT scores by the maximally obtained converted HAM-D score (i.e. a score of 53). Kernel centering (standard scaling) was used for scaling of MRI features. All machine learning analyses were implemented using the scikit-learn toolbox (v0.19.1) for Python (v2.7.15) (15). The primary performance metric was the area under the receiver operator characteristic (AUC) curve and reported metrics were averaged across cross-validation iterations (16). Balanced accuracy (average of sensitivity and specificity), sensitivity, specificity, positive predictive value (PPV) and negative predictive value (NPV) are reported in Supplementary Tables. Statistical significance of model performance was assessed using a label permutation-testing framework, in which the whole classification procedure was repeated 1000 times while randomly permuting the labels of the classes so that the labels no longer matched the real data in any meaningful way, in order to obtain empirical null-distribution for statistical testing (17). Label permutation of the classes was performed across centers for CV with internal validation, and permuted within centers for external validation (LOSO).

95% confidence intervals (CI) for AUC were computed using the modified Wald-method (18). It should be noted that the 95% CI were to be considered illustrative only. To reduce computational burden, only spatial ICA classifications that resulted in AUC>0.75 for either treatment remission or response were tested for significance with permutations. Finally, we assessed classification performance for multi-modal classifications by combining anatomical and functional features through feature concatenation: namely regional neuromorphometrics GM volumes with either ICA or Power-atlas based FC, and voxel-wise GM with either ICA or Power-atlas based FC. Obtained p-values were corrected for multiple comparisons using False Discovery Rate (FDR; two-stage (non-negative); alpha=0.05). FDR correction was applied separately for classification results obtained using either internal or external validation; the full dataset or three largest centers only; and for unimodal, multimodal and individual ICA spatial components, leading to 12 distinct families with qFDR set to (0.05/12=)0.00417. Missing p-values for ICA components that did not result in >0.75 AUC classification were filled in with ones for conservative FDR correction.

**Anatomical localization**

To investigate which regions contributed most to the SVM classification for the voxel-wise whole brain approach, the methods of Gaonkara *et al.* were used to derive p-values for the weights assigned by the classifier (19). In this approach a statistic was computed that was a combination of the weight component value and the size of the margin, providing a better metric than looking into the weight values only. An analytical approximation to the null-distribution obtained through permutation tests was used to calculate p-values. Previous work had shown that this approximation showed good correlation with empirical permutation tests (19). We only reported p-value feature importances for voxel-wise GM classifications for anatomical localization.

**Learning curves**

To evaluate the relationship between the size of the training set and classification performance, we computed learning curves for which we assessed classification performance using different proportions of training data. Learning curves were only computed for our best performing unimodal models, namely classification of treatment remission using GM data, as well as for our best performing multimodal model, using a combination of GM and ICA-based FC, both derived from data from the three largest centers with internal validation (**Supplementary Figure 2**). For both models, we performed classifications using 10% to 80% of training data (with increments of 10%) and 20% testing data with 10 CV splits, and repeated this procedure 100 times per proportion of training data used. The learning curves depict the average AUC obtained across the 100 iterations per proportion training data used. Average classification accuracy reached 0.83 and 0.84 AUC for unimodal and multimodal classifiers respectively, with AUC>0.75 for all resamplings at 50% of the data (N=55). The AUC was higher than 0.8 for all resamplings at 60% of the data (N=66). Both learning curves did not appear saturated, suggesting that model performance could still increase when using larger training samples.

### **S2 Supplementary Figures**

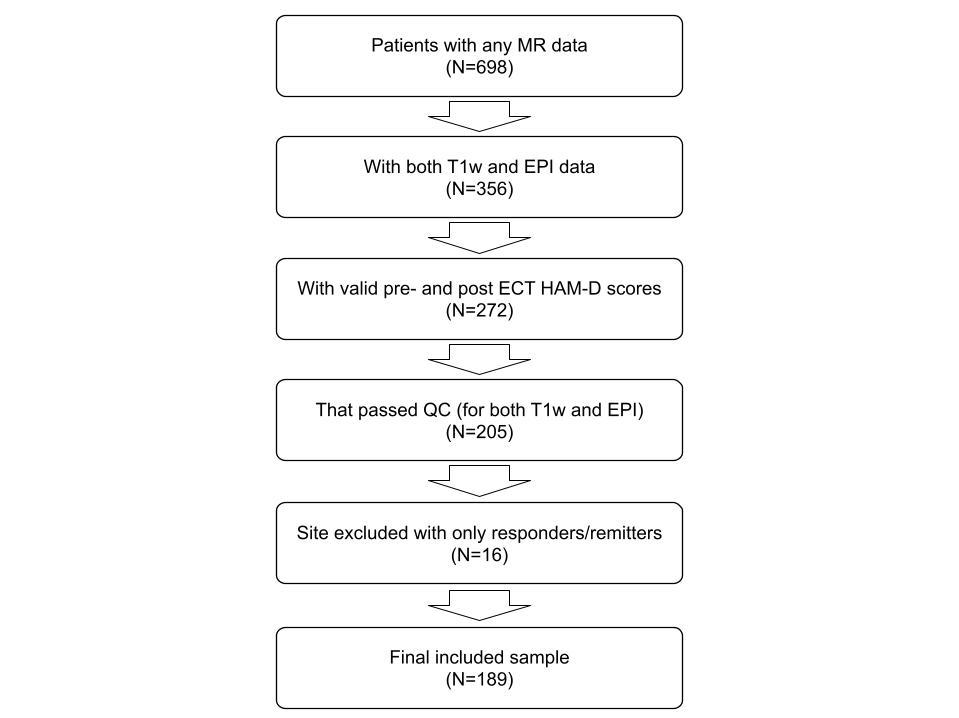

**Supplementary Figure 1.** Flowchart of patients included for analysis in this study. QC=quality control

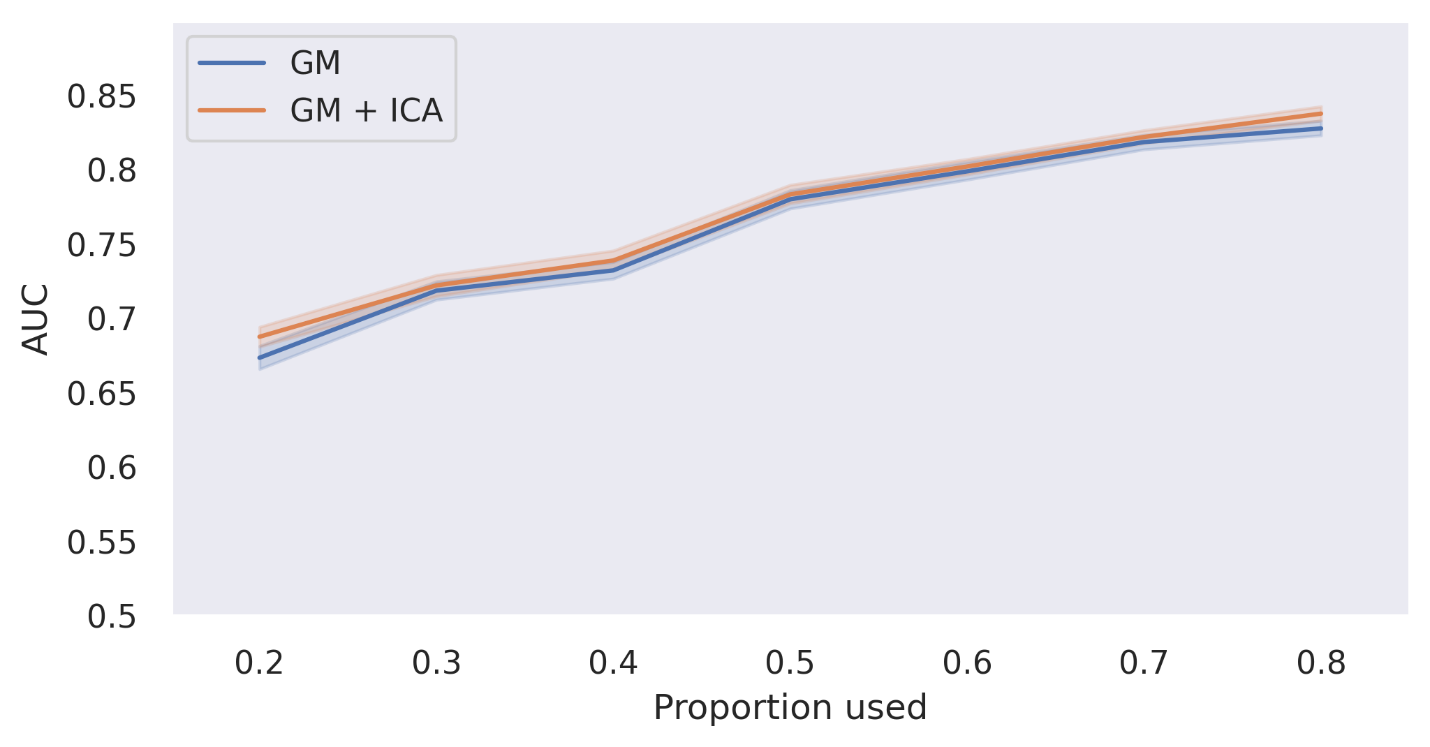

**Supplementary Figure 2.** Learning curves for best performing unimodal model (using voxel-wise gray matter volumes) and multimodal model (using voxel-wise gray matter combined with ICA-based FC) for remission classification using data from the three largest samples. The plotted line depicts the average AUC obtained for classifications using different proportions of training data (using 20% up to 80% of the data, with increments of 10% and 100 iterations per proportion used). Outer lines depict 95% confidence intervals. GM = voxel-wise gray matter volumes, ICA = ICA-based functional connectivity

### **S3 Supplementary Tables**

| **Center** | **BP** | **MDD single episode** | **MDD recurrent** | **Total** |
| --- | --- | --- | --- | --- |
| 1 | 7 | 0 | 35 | 42 |
| 2 | 0 | 2 | 27 | 29 |
| 3 | 0 | 4 | 11 | 15 |
| 4 | 2 | 0 | 12 | 14 |
| 5a* | 0 | 1 | 18 | 19 |
| 5b* | 8 | 3 | 27 | 38 |
| 6 | 5 | 0 | 13 | 18 |
| 7 | 0 | 4 | 10 | 14 |
| **Total** | 22 | 14 | 153 | 189 |

**Supplementary Table 1.** Diagnoses per center according to ICD-10. Asterisks depict diagnoses for the two samples obtained from center 5 with different inclusion criteria. BP=Bipolar Disorder, MDD=Major Depressive Disorder.

| **Center** | **Tesla** | **TR (ms)** | **TE (ms)** | **Flip Angle** | **Voxel-size (mm)** |
| --- | --- | --- | --- | --- | --- |
| 1 | 3 | 2530 | 1.74-7.32 | 7 | 1.00 x 1.00 x 1.30 |
| 2 | 3 | 2530 | 1.64-9.08 | 7 | 1.00 x 1.00 x 1.00 |
| 3 | 3 | 7840 | 3.02 | 12 | 0.94 x 0.94 x 1.00 |
| 4 | 3 | 7830 | 3.02 | 8 | 0.94 x 0.94 x 1.00 |
| 5 | 1.5 | 7700 | 3.50 | 15 | 1.07 x 1.07 x 1.10 |
| 6 | 3 | 2530 | 1.69 | 7 | 1.00 x 1.00 x 1.00 |
| 7 | 1.5 | 2730 | 2.95 | 7 | 1.00 x 1.00 x 1.00 |

**Supplementary Table 2.** MRI acquisition parameters used to obtain structural data for the different centers. More info on MRI acquisition is found elsewhere (20).

| **Center** | **Tesla** | **TR (ms)** | **TE (ms)** | **Flip Angle** | **Voxel-size (mm)** | **Volumes** |
| --- | --- | --- | --- | --- | --- | --- |
| 1 | 3 | 2000 | 30 | 70 | 3.44 x 3.44 x 5.00 | 180 |
| 2 | 3 | 2000 | 29 | 75 | 3.75 x 3.75 x 4.55 | 165 |
| 3 | 3 | 1800 | 35 | 80 | 3.30 x 3.30 x 3.30 | 202 |
| 4 | 3 | 2000 | 30 | 77 | 3.75 x 3.75 x 3.00 | 150 |
| 5 | 1.5 | 1868 | 30 | 90 | 2.40 x 2.40 x 4.50 | 150 |
| 6 | 3 | 3000 | 30 | 85 | 3.00 x 3.00 x 3.00 | 124 |
| 7 | 1.5 | 1870 | 35 | 80 | 3.50 x 3.50 x 3.50 | 266 |

**Supplementary Table 3.** MRI acquisition parameters used to obtain functional resting-state data for the different centers. More info on MRI acquisition is found elsewhere (20).

|  | **Total sample (n=109)** | | **Responders (n=74)** | | **Non-responders (n=35)** | | **R vs NR** |
| --- | --- | --- | --- | --- | --- | --- | --- |
|  | mean | std | mean | std | mean | std | p |
| Age | 51.9 | 15.3 | 54.7 | 12.9 | 46.0 | 17.7 | 0.013* |
| Sex (m/f) | 46/63 | n.a. | 32/42 | n.a. | 14/21 | n.a. | 0.91 |
| Laterality (RUL/BL; n=108) | 100/8 | n.a. | 67/7 | n.a. | 33/1 | n.a. | 0.42 |
| HAM-D pre-treatment | 25.7 | 6.9 | 26.6 | 7.1 | 23.9 | 7.4 | 0.046* |
| HAM-D post-treatment | 9.5 | 7.8 | 5.3 | 4.3 | 18.5 | 5.4 | 8.41E-18* |
| HAM-D change | 16.2 | 10.2 | 21.3 | 7.6 | 5.4 | 5.3 | 1.35E-21* |
| Diagnosis (UP/BP) | 104/15 | n.a. | 64/10 | n.a. | 30/5 | n.a. | 0.85 |
| Total ECT sessions | 13.5 | 6.1 | 13.4 | 6.4 | 13.6 | 5.4 | 0.88 |

**Supplementary Table 4.** Demographics of three largest centers used for analyses, with subject demographics and comparisons between ECT responders and non-responders. Abbreviations: R vs NR: responders versus non-responders; m: male; f: female; RUL: right unilateral ECT initially, BL: bilateral ECT initially; HAM-D: Hamilton Rating scale for depression; UP: unipolar depression; BP: bipolar depression. Asterisks depict significance using independent t-test or chi-squared test.

|  | **Total sample (n=109)** | | **Remitters (n=51)** | | **Non-remitters (n=58)** | | **R vs NR** |
| --- | --- | --- | --- | --- | --- | --- | --- |
|  | mean | std | mean | std | mean | std | p |
| Age | 51.9 | 15.3 | 56.9 | 13.1 | 47.5 | 15.6 | 0.001* |
| Sex (m/f) | 83/106 | n.a. | 18/33 | n.a. | 28/30 | n.a. | 0.24 |
| Laterality (RUL/BL; n=108) | 100/8 | n.a. | 44/7 | n.a. | 56/1 | n.a. | 0.045* |
| HAM-D pre-treatment | 25.7 | 6.9 | 26.6 | 7.4 | 24.9 | 6.3 | 0.21 |
| HAM-D post-treatment | 9.5 | 7.8 | 2.9 | 2.3 | 15.3 | 6.0 | 2.16E-23* |
| HAM-D change | 16.2 | 10.2 | 23.7 | 7.5 | 9.6 | 7.2 | 1.43E-16* |
| Diagnosis (UP/BP) | 104/15 | n.a. | 45/6 | n.a. | 49/9 | n.a. | 0.77 |
| Total ECT sessions | 13.5 | 6.1 | 13.4 | 6.5 | 13.4 | 5.7 | 0.83 |

**Supplementary Table 5.** Demographics of three largest centers used for analyses, with subject demographics and comparisons between ECT remitters and non-remitters. Abbreviations: R vs NR: remitters versus non-remitters; m: male; f: female; RUL: right unilateral ECT initially, BL: bilateral ECT initially; HAM-D: Hamilton Rating scale for depression; UP: unipolar depression; BP: bipolar depression. Asterisks depict significance using independent t-test or chi-squared test.

| **Validation** | **Modality** | **Classification** | **AUC** (95% CI) | **p-value**  (FDR corrected) | **Balanced Accuracy** | **Sensitivity** | **Specificity** | **PPV** | **NPV** |
| --- | --- | --- | --- | --- | --- | --- | --- | --- | --- |
| internal | baseline | Remission | 0.620* (0.540 - 0.700) | 0.02597 | 0.569 | 0.639 | 0.498 | 0.452 | 0.673 |
| internal | biobank_gica_comps_c42 | Remission | 0.700 (0.624 - 0.776) | 0.20779 | 0.637 | 0.740 | 0.533 | 0.517 | 0.777 |
| internal | biobank_gica_comps_c52 | Remission | 0.599 (0.518 - 0.680) | 1.00000 | 0.598 | 0.811 | 0.385 | 0.462 | 0.774 |
| internal | GICA-DR FC | Remission | 0.669* (0.591 - 0.747) | 0.00500 | 0.580 | 0.687 | 0.472 | 0.501 | 0.491 |
| internal | VBM | Remission | 0.628* (0.548 - 0.708) | 0.01249 | 0.586 | 0.558 | 0.613 | 0.490 | 0.687 |
| internal | neuromorphometrics | Remission | 0.577 (0.495 - 0.658) | 0.08991 | 0.529 | 0.522 | 0.536 | 0.418 | 0.586 |
| internal | Power FC | Remission | 0.656* (0.578 - 0.735) | 0.00999 | 0.595 | 0.584 | 0.605 | 0.510 | 0.702 |
| external | baseline | Remission | 0.507 (0.425 - 0.590) | 0.65078 | 0.469 | 0.420 | 0.519 | 0.336 | 0.579 |
| external | biobank_gica_comps_c42 | Remission | 0.629 (0.549 - 0.709) | 1.00000 | 0.524 | 0.684 | 0.365 | 0.443 | 0.726 |
| external | biobank_gica_comps_c52 | Remission | 0.520 (0.438 - 0.603) | 1.00000 | 0.503 | 0.880 | 0.126 | 0.381 | 0.288 |
| external | GICA-DR FC | Remission | 0.512 (0.429 - 0.594) | 0.65078 | 0.475 | 0.233 | 0.718 | 0.130 | 0.632 |
| external | VBM | Remission | 0.513 (0.430 - 0.595) | 0.65078 | 0.518 | 0.480 | 0.556 | 0.407 | 0.637 |
| external | neuromorphometrics | Remission | 0.576 (0.494 - 0.657) | 0.57942 | 0.537 | 0.427 | 0.647 | 0.315 | 0.677 |
| external | Power FC | Remission | 0.575 (0.493 - 0.656) | 0.57942 | 0.542 | 0.516 | 0.568 | 0.525 | 0.622 |

**Supplementary Table 6.** Unimodal remission classification using all centers. Asterisks for AUC depict statistical significance (qFDR=0.05/12) following permutation testing with multiple comparison correction. Biobank_gica_comps_c42 is a network centered around the temporal lobes, biobank_gica_comps_c52 is a network located in frontopolar cortex. Abbreviations: AUC: area under the receiver operator characteristic curve; CI: confidence intervals; FDR: two-stage False Discovery Rate; PPV: positive predictive value; NPV: negative predictive value.

| **Validation** | **Modality** | **Classification** | **AUC** (95% CI) | **p-value**  (FDR corrected) | **Balanced Accuracy** | **Sensitivity** | **Specificity** | **PPV** | **NPV** |
| --- | --- | --- | --- | --- | --- | --- | --- | --- | --- |
| internal | VBM + ICA-DR FC | Remission | 0.640* (0.561 - 0.719) | 0.01349 | 0.598 | 0.565 | 0.630 | 0.508 | 0.696 |
| internal | VBM + Power FC | Remission | 0.673* (0.596 - 0.751) | 0.00300 | 0.636 | 0.557 | 0.715 | 0.571 | 0.716 |
| internal | NMM + ICA-DR FC | Remission | 0.642* (0.563 - 0.721) | 0.01349 | 0.600 | 0.571 | 0.630 | 0.511 | 0.693 |
| internal | NMM + Power FC | Remission | 0.676* (0.599 - 0.754) | 0.00300 | 0.619 | 0.624 | 0.613 | 0.523 | 0.722 |
| external | VBM + ICA-DR FC | Remission | 0.517 (0.434 - 0.599) | 0.58475 | 0.493 | 0.367 | 0.620 | 0.372 | 0.606 |
| external | VBM + Power FC | Remission | 0.566 (0.484 - 0.648) | 0.57542 | 0.541 | 0.337 | 0.746 | 0.428 | 0.647 |
| external | NMM + ICA-DR FC | Remission | 0.514 (0.432 - 0.597) | 0.58475 | 0.490 | 0.352 | 0.627 | 0.387 | 0.561 |
| external | NMM + Power FC | Remission | 0.635 (0.556 - 0.715) | 0.06394 | 0.526 | 0.455 | 0.597 | 0.492 | 0.562 |

**Supplementary Table 7.** Multimodal remission classification using all centers. Asterisks for AUC depict statistical significance (qFDR=0.05/12) following permutation testing with multiple comparison correction. Abbreviations: AUC: area under the receiver operator characteristic curve; CI: confidence intervals; FDR: two-stage False Discovery Rate; PPV: positive predictive value; NPV: negative predictive value.

| **Validation** | **Modality** | **Classification** | **AUC** (95% CI) | **p-value**  (FDR corrected) | **Balanced Accuracy** | **Sensitivity** | **Specificity** | **PPV** | **NPV** |
| --- | --- | --- | --- | --- | --- | --- | --- | --- | --- |
| internal | baseline | Remission | 0.648* (0.544 - 0.752) | 0.02747 | 0.569 | 0.688 | 0.449 | 0.574 | 0.529 |
| internal | biobank_gica_comps_c42 | Remission | 0.751 (0.657 - 0.845) | 0.05195 | 0.686 | 0.740 | 0.632 | 0.677 | 0.724 |
| internal | biobank_gica_comps_c52 | Remission | 0.797 (0.710 - 0.885) | 0.05195 | 0.769 | 0.819 | 0.719 | 0.747 | 0.809 |
| internal | GICA-DR FC | Remission | 0.741* (0.645 - 0.836) | 0.00999 | 0.588 | 0.863 | 0.313 | 0.578 | 0.372 |
| internal | VBM | Remission | 0.825* (0.743 - 0.908) | 0.00999 | 0.726 | 0.737 | 0.714 | 0.744 | 0.751 |
| internal | neuromorphometrics | Remission | 0.524 (0.415 - 0.632) | 0.44622 | 0.511 | 0.563 | 0.458 | 0.503 | 0.364 |
| internal | Power FC | Remission | 0.690* (0.589 - 0.791) | 0.01665 | 0.616 | 0.730 | 0.503 | 0.606 | 0.646 |
| external | baseline | Remission | 0.473 (0.364 - 0.581) | 0.79545 | 0.501 | 0.870 | 0.133 | 0.503 | 0.133 |
| external | biobank_gica_comps_c42 | Remission | 0.627 (0.521 - 0.732) | 1.00000 | 0.546 | 0.636 | 0.456 | 0.318 | 0.632 |
| external | biobank_gica_comps_c52 | Remission | 0.560 (0.452 - 0.669) | 1.00000 | 0.556 | 0.755 | 0.358 | 0.531 | 0.574 |
| external | GICA-DR FC | Remission | 0.587 (0.480 - 0.694) | 0.49950 | 0.477 | 0.333 | 0.620 | 0.253 | 0.411 |
| external | VBM | Remission | 0.697 (0.597 - 0.798) | 0.08991 | 0.565 | 0.317 | 0.814 | 0.685 | 0.531 |
| external | neuromorphometrics | Remission | 0.481 (0.372 - 0.590) | 0.79545 | 0.477 | 0.071 | 0.884 | 0.222 | 0.487 |
| external | Power FC | Remission | 0.581 (0.473 - 0.688) | 0.49950 | 0.478 | 0.561 | 0.395 | 0.458 | 0.528 |

**Supplementary Table 8.** Unimodal remission classification using three largest centers. Asterisks for AUC depict statistical significance (qFDR=0.05/12) following permutation testing with multiple comparison correction. Biobank_gica_comps_c42 is a network centered around the temporal lobes, biobank_gica_comps_c52 is a network located in frontopolar cortex. Abbreviations: AUC: area under the receiver operator characteristic curve; CI: confidence intervals; FDR: two-stage False Discovery Rate; PPV: positive predictive value; NPV: negative predictive value.

| **Validation** | **Modality** | **Classification** | **AUC** (95% CI) | **p-value**  (FDR corrected) | **Balanced Accuracy** | **Sensitivity** | **Specificity** | **PPV** | **NPV** |
| --- | --- | --- | --- | --- | --- | --- | --- | --- | --- |
| internal | VBM + ICA-DR FC | Remission | 0.834* (0.753 - 0.915) | 0.00167 | 0.727 | 0.721 | 0.734 | 0.752 | 0.741 |
| internal | VBM + Power FC | Remission | 0.817* (0.732 - 0.901) | 0.00167 | 0.734 | 0.715 | 0.753 | 0.762 | 0.736 |
| internal | NMM + ICA-DR FC | Remission | 0.746* (0.651 - 0.841) | 0.00167 | 0.683 | 0.657 | 0.709 | 0.712 | 0.686 |
| internal | NMM + Power FC | Remission | 0.665* (0.562 - 0.768) | 0.00599 | 0.598 | 0.639 | 0.557 | 0.603 | 0.613 |
| external | VBM + ICA-DR FC | Remission | 0.698* (0.599 - 0.798) | 0.02797 | 0.553 | 0.305 | 0.801 | 0.608 | 0.529 |
| external | VBM + Power FC | Remission | 0.733* (0.636 - 0.829) | 0.02797 | 0.672 | 0.592 | 0.752 | 0.646 | 0.607 |
| external | NMM + ICA-DR FC | Remission | 0.514 (0.406 - 0.623) | 0.57200 | 0.449 | 0.273 | 0.624 | 0.457 | 0.476 |
| external | NMM + Power FC | Remission | 0.550 (0.442 - 0.659) | 0.57200 | 0.530 | 0.439 | 0.621 | 0.524 | 0.533 |

**Supplementary Table 9.** Multimodal remission classification using three largest centers. Asterisks for AUC depict statistical significance (qFDR=0.05/12) following permutation testing with multiple comparison correction. Abbreviations: AUC: area under the receiver operator characteristic curve; CI: confidence intervals; FDR: two-stage False Discovery Rate; PPV: positive predictive value; NPV: negative predictive value.

| **Validation** | **Modality** | **Classification** | **AUC** (95% CI) | **p-value**  (FDR corrected) | **Balanced Accuracy** | **Sensitivity** | **Specificity** | **PPV** | **NPV** |
| --- | --- | --- | --- | --- | --- | --- | --- | --- | --- |
| internal | baseline | Response | 0.640* (0.561 - 0.719) | 0.00500 | 0.602 | 0.822 | 0.382 | 0.644 | 0.673 |
| internal | biobank_gica_comps_c42 | Response | 0.620 (0.540 - 0.700) | 1.00000 | 0.573 | 0.634 | 0.511 | 0.643 | 0.522 |
| internal | biobank_gica_comps_c52 | Response | 0.513 (0.430 - 0.595) | 1.00000 | 0.505 | 0.633 | 0.376 | 0.567 | 0.442 |
| internal | GICA-DR FC | Response | 0.558 (0.476 - 0.640) | 0.25863 | 0.511 | 0.515 | 0.508 | 0.512 | 0.418 |
| internal | VBM | Response | 0.565 (0.483 - 0.647) | 0.16109 | 0.525 | 0.545 | 0.506 | 0.605 | 0.457 |
| internal | neuromorphometrics | Response | 0.470 (0.388 - 0.553) | 0.79820 | 0.479 | 0.508 | 0.450 | 0.542 | 0.372 |
| internal | Power FC | Response | 0.596 (0.515 - 0.677) | 0.08991 | 0.540 | 0.500 | 0.579 | 0.641 | 0.446 |
| external | baseline | Response | 0.548 (0.465 - 0.630) | 0.65078 | 0.500 | 0.752 | 0.249 | 0.605 | 0.386 |
| external | biobank_gica_comps_c42 | Response | 0.563 (0.482 - 0.645) | 1.00000 | 0.538 | 0.737 | 0.339 | 0.614 | 0.445 |
| external | biobank_gica_comps_c52 | Response | 0.425 (0.343 - 0.506) | 1.00000 | 0.462 | 0.789 | 0.135 | 0.486 | 0.192 |
| external | GICA-DR FC | Response | 0.463 (0.381 - 0.545) | 0.79920 | 0.527 | 0.328 | 0.725 | 0.478 | 0.401 |
| external | VBM | Response | 0.487 (0.405 - 0.570) | 0.71678 | 0.467 | 0.466 | 0.468 | 0.538 | 0.378 |
| external | neuromorphometrics | Response | 0.426 (0.344 - 0.508) | 0.85714 | 0.445 | 0.399 | 0.491 | 0.454 | 0.306 |
| external | Power FC | Response | 0.524 (0.442 - 0.606) | 0.65078 | 0.518 | 0.496 | 0.541 | 0.611 | 0.305 |

**Supplementary Table 10.** Unimodal response classification using all centers. Asterisks for AUC depict statistical significance (qFDR=0.05/12) following permutation testing with multiple comparison correction. Biobank_gica_comps_c42 is a network centered around the temporal lobes, biobank_gica_comps_c52 is a network located in frontopolar cortex. Abbreviations: AUC: area under the receiver operator characteristic curve; CI: confidence intervals; FDR: two-stage False Discovery Rate; PPV: positive predictive value; NPV: negative predictive value.

| **Validation** | **Modality** | **Classification** | **AUC** (95% CI) | **p-value**  (FDR corrected) | **Balanced Accuracy** | **Sensitivity** | **Specificity** | **PPV** | **NPV** |
| --- | --- | --- | --- | --- | --- | --- | --- | --- | --- |
| internal | VBM + ICA-DR FC | Response | 0.564 (0.482 - 0.646) | 0.13444 | 0.524 | 0.543 | 0.505 | 0.604 | 0.453 |
| internal | VBM + Power FC | Response | 0.583 (0.502 - 0.665) | 0.11389 | 0.541 | 0.541 | 0.542 | 0.626 | 0.472 |
| internal | NMM + ICA-DR FC | Response | 0.556 (0.474 - 0.638) | 0.13936 | 0.515 | 0.521 | 0.508 | 0.538 | 0.446 |
| internal | NMM + Power FC | Response | 0.570 (0.488 - 0.651) | 0.13444 | 0.527 | 0.486 | 0.568 | 0.630 | 0.435 |
| external | VBM + ICA-DR FC | Response | 0.490 (0.407 - 0.572) | 0.61439 | 0.456 | 0.445 | 0.468 | 0.512 | 0.375 |
| external | VBM + Power FC | Response | 0.479 (0.397 - 0.562) | 0.61439 | 0.473 | 0.485 | 0.462 | 0.481 | 0.345 |
| external | NMM + ICA-DR FC | Response | 0.511 (0.428 - 0.593) | 0.58475 | 0.465 | 0.546 | 0.383 | 0.566 | 0.315 |
| external | NMM + Power FC | Response | 0.526 (0.444 - 0.608) | 0.58475 | 0.506 | 0.357 | 0.655 | 0.600 | 0.327 |

**Supplementary Table 11.** Multimodal response classification using all centers. Asterisks for AUC depict statistical significance (qFDR=0.05/12) following permutation testing with multiple comparison correction. Abbreviations: AUC: area under the receiver operator characteristic curve; CI: confidence intervals; FDR: two-stage False Discovery Rate; PPV: positive predictive value; NPV: negative predictive value.

| **Validation** | **Modality** | **Classification** | **AUC** (95% CI) | **p-value**  (FDR corrected) | **Balanced Accuracy** | **Sensitivity** | **Specificity** | **PPV** | **NPV** |
| --- | --- | --- | --- | --- | --- | --- | --- | --- | --- |
| internal | baseline | Response | 0.653* (0.550 - 0.757) | 0.03596 | 0.628 | 0.845 | 0.411 | 0.755 | 0.585 |
| internal | biobank_gica_comps_c42 | Response | 0.597 (0.490 - 0.704) | 1.00000 | 0.549 | 0.600 | 0.497 | 0.719 | 0.386 |
| internal | biobank_gica_comps_c52 | Response | 0.687 (0.586 - 0.788) | 0.31169 | 0.621 | 0.631 | 0.610 | 0.784 | 0.444 |
| internal | GICA-DR FC | Response | 0.659* (0.556 - 0.763) | 0.04710 | 0.602 | 0.717 | 0.486 | 0.736 | 0.465 |
| internal | VBM | Response | 0.636* (0.531 - 0.741) | 0.03996 | 0.587 | 0.664 | 0.510 | 0.750 | 0.438 |
| internal | neuromorphometrics | Response | 0.495 (0.386 - 0.604) | 0.56643 | 0.499 | 0.453 | 0.544 | 0.670 | 0.287 |
| internal | Power FC | Response | 0.637* (0.532 - 0.741) | 0.04870 | 0.596 | 0.491 | 0.701 | 0.793 | 0.382 |
| external | baseline | Response | 0.444 (0.336 - 0.552) | 0.81141 | 0.513 | 0.655 | 0.370 | 0.547 | 0.341 |
| external | biobank_gica_comps_c42 | Response | 0.528 (0.419 - 0.637) | 1.00000 | 0.466 | 0.393 | 0.538 | 0.433 | 0.293 |
| external | biobank_gica_comps_c52 | Response | 0.538 (0.430 - 0.647) | 1.00000 | 0.504 | 0.580 | 0.429 | 0.703 | 0.304 |
| external | GICA-DR FC | Response | 0.504 (0.396 - 0.613) | 0.79545 | 0.451 | 0.731 | 0.172 | 0.679 | 0.229 |
| external | VBM | Response | 0.514 (0.406 - 0.623) | 0.79545 | 0.513 | 0.667 | 0.360 | 0.706 | 0.390 |
| external | neuromorphometrics | Response | 0.380 (0.274 - 0.486) | 0.92807 | 0.458 | 0.507 | 0.409 | 0.654 | 0.202 |
| external | Power FC | Response | 0.553 (0.445 - 0.661) | 0.66184 | 0.508 | 0.637 | 0.379 | 0.702 | 0.224 |

**Supplementary Table 12.** Unimodal response classification using three largest centers. Asterisks for AUC depict statistical significance (qFDR=0.05/12) following permutation testing with multiple comparison correction. Biobank_gica_comps_c42 is a network centered around the temporal lobes, biobank_gica_comps_c52 is a network located in frontopolar cortex. Abbreviations: AUC: area under the receiver operator characteristic curve; CI: confidence intervals; FDR: two-stage False Discovery Rate; PPV: positive predictive value; NPV: negative predictive value.

| **Validation** | **Modality** | **Classification** | **AUC** (95% CI) | **p-value**  (FDR corrected) | **Balanced Accuracy** | **Sensitivity** | **Specificity** | **PPV** | **NPV** |
| --- | --- | --- | --- | --- | --- | --- | --- | --- | --- |
| internal | VBM + ICA-DR FC | Response | 0.650* (0.546 - 0.754) | 0.02831 | 0.594 | 0.660 | 0.527 | 0.760 | 0.439 |
| internal | VBM + Power FC | Response | 0.686* (0.585 - 0.788) | 0.00599 | 0.640 | 0.662 | 0.619 | 0.796 | 0.487 |
| internal | NMM + ICA-DR FC | Response | 0.628* (0.523 - 0.733) | 0.04567 | 0.590 | 0.689 | 0.491 | 0.745 | 0.444 |
| internal | NMM + Power FC | Response | 0.609 (0.503 - 0.715) | 0.05807 | 0.558 | 0.455 | 0.661 | 0.727 | 0.359 |
| external | VBM + ICA-DR FC | Response | 0.500 (0.391 - 0.609) | 0.57200 | 0.533 | 0.707 | 0.360 | 0.719 | 0.397 |
| external | VBM + Power FC | Response | 0.529 (0.420 - 0.638) | 0.57200 | 0.508 | 0.534 | 0.482 | 0.692 | 0.331 |
| external | NMM + ICA-DR FC | Response | 0.463 (0.354 - 0.572) | 0.65435 | 0.467 | 0.508 | 0.427 | 0.688 | 0.272 |
| external | NMM + Power FC | Response | 0.554 (0.446 - 0.662) | 0.57200 | 0.551 | 0.435 | 0.667 | 0.919 | 0.241 |

**Supplementary Table 13.** Multimodal response classification using three largest centers. Asterisks for AUC depict statistical significance (qFDR=0.05/12) following permutation testing with multiple comparison correction. Abbreviations: AUC: area under the receiver operator characteristic curve; CI: confidence intervals; FDR: two-stage False Discovery Rate; PPV: positive predictive value; NPV: negative predictive value.
